# Effectiveness of Computer-Aided interventions and Virtual Reality in Wrist and Hand rehabilitation: a Scoping Review Protocol

**DOI:** 10.1101/2025.01.28.25320373

**Authors:** Nicola Tonial, Valentina Scalise, Filippo Maselli, Elena Benedettini, Michela Pili, Sheila Santandrea

## Abstract

**Background:** To date, advances in the technological and medical fields have increased the application of virtual reality tools, smartphone apps, and computer-aided therapy in clinical and rehabilitation settings. These tools have been shown to enhance patient involvement and improve self-efficacy in their treatment pathways. However, in orthopaedic and rheumatology fields, studies remain limited, and comprehensive reviews providing an exhaustive overview are lacking.

**Objective:** The purpose of this Scoping Review is to provide a general overview of the use of virtual reality and computer aided equipment in musculoskeletal and rheumatological rehabilitation of the wrist and hand, outlining the main types of instruments used, how they are applied, describing any gaps in the literature and possible future developments on the use of VR and PC-aided treatments.

**Eligibility criteria:** This study will contain articles that included a population with hand and/or wrist deficits attributable to musculoskeletal and/or rheumatological pathologies or traumas and who undertook a rehabilitation programme focusing on virtual reality or computer aided devices. All studies with subjects suffering from neurological pathologies, such as stroke or Parkinson’s disease, will be excluded.

**Methods:** This scoping review will be carried out following the guidance of the Joanna Briggs Institute (JBI). Articles will be searched in databases such as PubMed (Medline), Cochrane central, Scopus, Pedro and CINHALcomplete from December 2024. Additional studies will be selected through grey literature, such as Google Scholar. Article selection and data mining will be done by three reviewers, while a fourth will resolve any discrepancies or disputes. No restrictions on type of publication will apply; only articles written since 2014 and written in Italian or English will be considered.

**Ethics and dissemination:** Ethical approval is unnecessary for this study.

## Introduction

The complexity of the hand and wrist, encompassing tendons, bones, ligaments, and muscles, coupled with their vital roles in daily life, makes rehabilitation after trauma, pathology, or surgery particularly challenging ^(1)^. Delayed treatments, often due to physiotherapists’ or patients’ underestimation of symptoms, can significantly hinder recovery^(2)^. Fortunately, however, in recent years the treatment of injuries to this district has seen a major improvement thanks to the large number of studies that have investigated and described the biomechanics and functional anatomy of the hand and wrist, and thanks to new rehabilitation approach and surgical techniques that are increasingly sophisticated and precise ^(3)^.

After an injury, surgery or a period of immobilisation, such as a plaster cast, it is important to start physiotherapy as soon as possible. This is to ensure the best possible outcome^(4)^. The treatment should be helpful in reducing pain, improving the quality of movement, hand grip strength and range of motion, both of which are major problems after an immobilisation time, and which are essential in order to be able to involve the hand in daily life activities ^(4)^. During this rehabilitation pathway, however, the physiotherapist must consider four fundamental aspects for the success of patient-centred treatment: the number of physiotherapy treatments useful for the patient, the frequency and personalisation of treatment, continuous monitoring and objectification of the results achieved by the patient, and adherence to the rehabilitation treatment ^(5)^. And especially for the latter aspects, the use of Virtual Reality (VR) and applications via smartphone, tablet or PC appears to be a further effective means in clinical practice ^(6–8)^.

In recent years, interest in the use of VR in the medical community has increased significantly, due in part to its use in innovative rehabilitation treatments ^(9)^. VR is based on the principle of interaction between a user and an electronic device (e.g. a PC) that allows stimuli to be transmitted in real time ^(9)^. The patient can have the sensation of being in another place, the possibility of responding to stimuli or manipulating objects, as well as simulating real-life activities. All this in a safe and controlled manner ^(8,9)^. VR systems consist not only of input-output peripherals and specific software, but also of tools that provide visual, auditory and tactile feedback such as gloves, joysticks, etc., allowing a better experience and helping the patient to perform even potentially difficult and complex tasks in normal situations ^(9)^.

Different from VR is augmented reality (AR). Here, the real world is amplified by digital information, which can depend on the patient’s point of view in the physical world ^(10)^. This results in an interactive experience where a person perceives the information and, if desired, can modify it ^(10)^. The level of interaction in this type of reality goes from simple modifications of the physical perspective to the manipulation and eventual creation of new information ^(10)^.

The term ‘*Gamification*’ is defined as ‘the use of game design elements in non-game contexts’ ^(11)^. It aims to stimulate patient behaviour by improving engagement and interaction through more enjoyable and motivating activities. It also helps users experience positive emotions such as satisfaction and self-esteem, influencing the person’s attitude towards their health and well-being and improving adherence to treatment ^(11)^.

A large part of the world’s population also owns a smartphone or mobile device and uses it daily ^(12)^. It has been estimated, in fact, that around 500 million people worldwide use health apps on their mobile phones, and this number is constantly growing ^(12)^. For this reason, a possible modality of intervention may involve the use of these devices, both during but especially at the end of the rehabilitation process: for the patient, the execution of exercises wherever he or she wishes and at any time of day, and the possibility of communicating remotely with the therapist regarding symptoms or variations in the clinical picture are important aspects to be considered and make it possible to favour better self-efficacy and to improve adherence to treatment; for the therapist, the possibility of following the patient’s progress, defining remotely the objectives and treatment modalities most appropriate to the phases of the rehabilitation pathway reached make it possible to reduce waiting times and to treat more patients at the same time ^(6,8)^.

Unfortunately, despite the potential benefits, the high prevalence of musculoskeletal disorders and the need for evidence based physiotherapy, the study of rehabilitation through VR and technology in general in the musculoskeletal field remains under-researched compared to the literature in the neurological field ^(6,8,13)^.

### Objective

In accordance with the Joanna Briggs institute (JBI), scoping reviews have the task of outlining and defining key concepts, summarising the present knowledge and type of literature and highlighting any gaps in the literature ^(14)^. This scoping review aims to:

- give an exhaustive overview of studies concerning VR, mobile device applications and video games applied for the management of musculoskeletal and rheumatologic disorders in patients with wrist and/or hand problems;
- define the different types of tools, modes of application of VR and their tolerability in these subjects;
- highlight any gaps in the literature regarding virtual reality applied in the orthopaedic and rheumatology fields.

## Methods

The scoping review will follow JBI Methodology and include six steps: 1) identifying the research question, 2) identifying relevant studies, 3) study selection, 4) charting the data, 5) collating, summarising and reporting the results and eventually 6) consultation ^(15)^. The protocol was realised using the template proposed by Lely ^(16)^. The Preferred Reporting Item for Systematic Review and Meta Analysis extension for Scoping Reviews (PRISMA-SCR) will be used to complete this study ^(17)^. Finally, the protocol will be entered into *Medrxiv.org* for scientific approval.

### Review question

1. What is the current knowledge on VR interventions, smartphone apps and video games in physiotherapy, applied in patients with musculoskeletal and rheumatologic disorders in the wrist-hand region?
2. How is VR/AR/MR [mixed reality] applied in this district and which devices are most commonly used in the literature?
3. What advantages/disadvantages have been reported in the literature regarding the use of VR and other types of technology considered for hand and wrist rehabilitation?
4. Have any adverse events been reported during or after the use of these technologies?
5. Which outcome have been used in articles dealing with VR, mobile apps and video games for the musculoskeletal hand and wrist physiotherapy?
6. Are there particular geographical areas in the world that make greater use of this type of technology in the field of musculoskeletal and rheumatologic rehabilitation of the wrist and hand?
7. Are there any gaps or discrepancies in the literature concerning this topic?

## Eligibility criteria

Studies will be eligible for inclusion if they follow the Population, Concept and Context (PCC) framework criteria. Inclusion and exclusion criteria are reported on *Table 1*.

**Table 1:** Population, Concept and Context (PCC)

|  |  |  |
| --- | --- | --- |
|  | Inclusion criteria | Exclusion criteria |
| Population | Individuals with musculoskeletal/ rheumatologic hand and/or wrist deficits. No restriction of any age, gender, ethnicity. | Individuals with neurological disorders.<br>Animals.<br>Vitreous studies. |
| Concept | Devices that make use of augmented reality, mixed reality and virtual reality with different levels of immersion.<br>Apps for smartphones/tablets/PCs or other electronic devices and articles dealing with video games. | None |
| Context | Studies conducted in any setting, geographical area, social and cultural context. | None |

### Inclusion criteria

*Population*: individuals of all ages, genders and ethnicities, with any musculoskeletal or rheumatologic hand and/or wrist disorder diagnosed by any type of healthcare professional will be included.

*Concept*: This includes all devices that utilize AR, MR, and VR with varying levels of immersion. Apps for smartphones, tablets, PCs, or other electronic devices, as well as articles related to video games, will also be considered

*Context*: No restriction on the rehabilitation setting, cultural or social context and geographical area in which the articles were carried out will be applied.

*Sources*: This Scoping Review will consider both experimental and non-experimental articles, including non-randomised controlled trials, randomised controlled trials, observational studies, case reports, case-control studies and observational studies. Any systematic reviews that meet the requirements for inclusion in this study will also be included. Any article published since 2014 will be considered.

The decision to include studies published in the last 10 years is based on the rapid pace of technological evolution. Tools, devices, and applications developed more than a decade ago may no longer exist or adequately reflect the current state-of-the-art. By focusing on this time frame, we ensure a review of modern and clinically relevant tools that align with today’s needs, maintaining the relevance and validity of the findings.

Language: Any article written in English or Italian will be included.

### Exclusion Criteria

*Population*: Studies in vitro, in animals, or in a population of human subjects with neurologic disorders will not be taken into account.

All articles that do not meet the PCC framework described above will be excluded.

## Search strategy

The search strategy was carried out by 4 reviewers specialised in the fields of musculoskeletal and rheumatology rehabilitation. A preliminary search was carried out on Medline (via Pubmed) in order to identify key terms and to identify the articles most relevant to the topic under investigation. An example of a search string for the Pubmed database is presented in *Table 2*. A subsequent search will be carried out on the main clinical search engines such as: Pedro, Cochrane central, Scopus and CINAHL complete. In addition, a further search will involve reviewing grey literature and unpublished articles (letters to authors, books and sites such as Google Scholar and ResearchGate). The search strategy, including keywords (MESH terms) and free terms were repurposed and used in each database or information resource used.

**Table 2:**
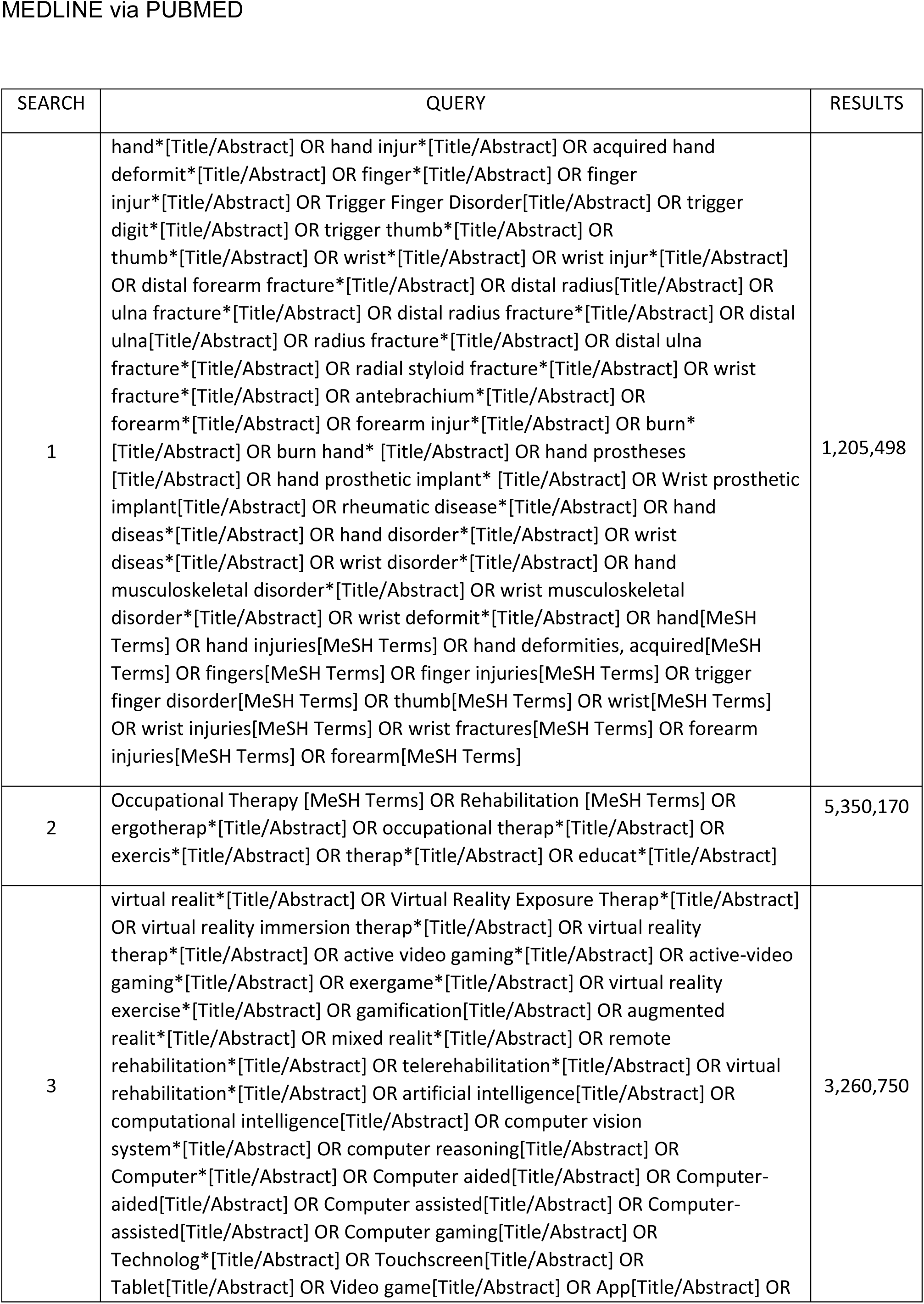
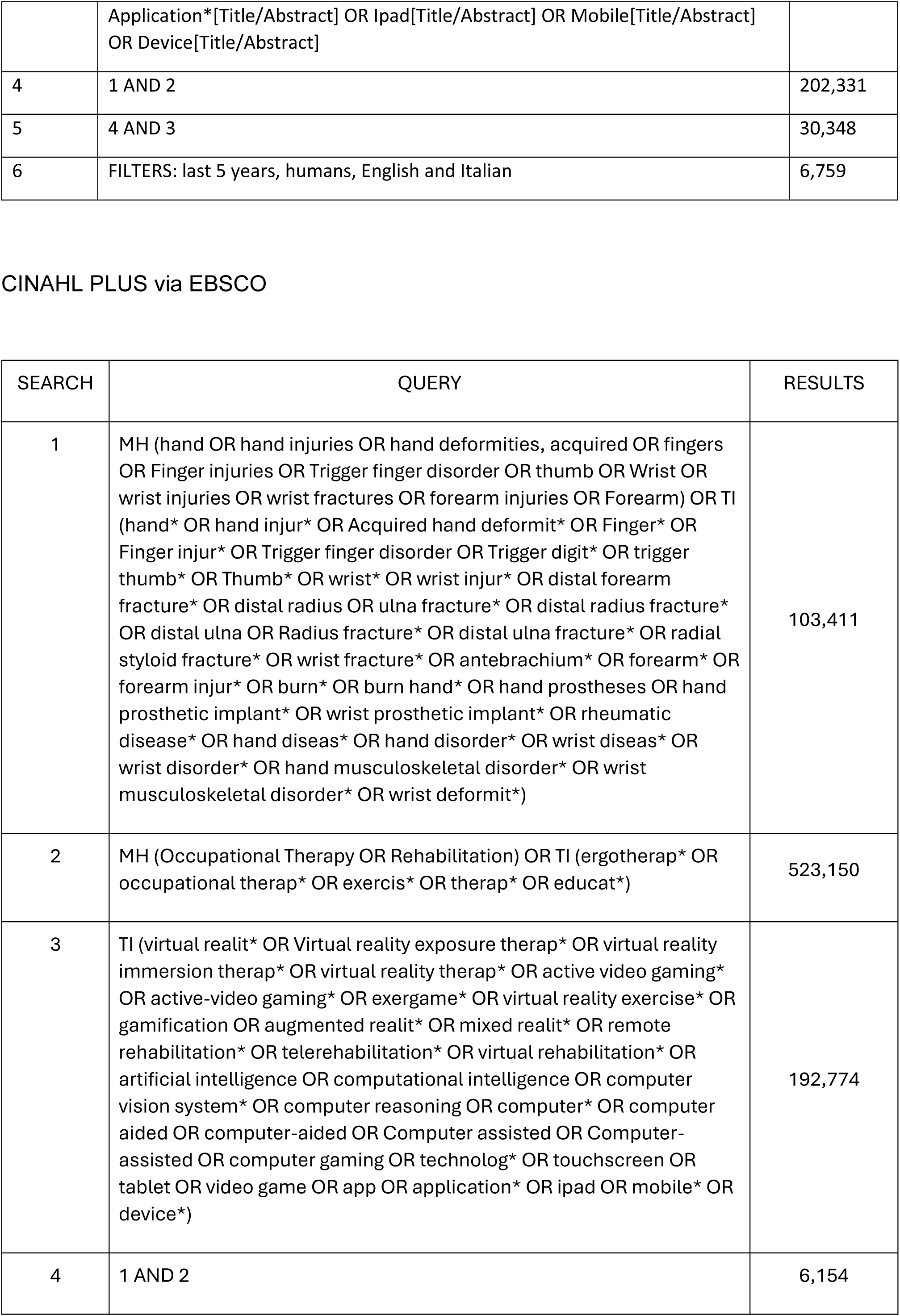
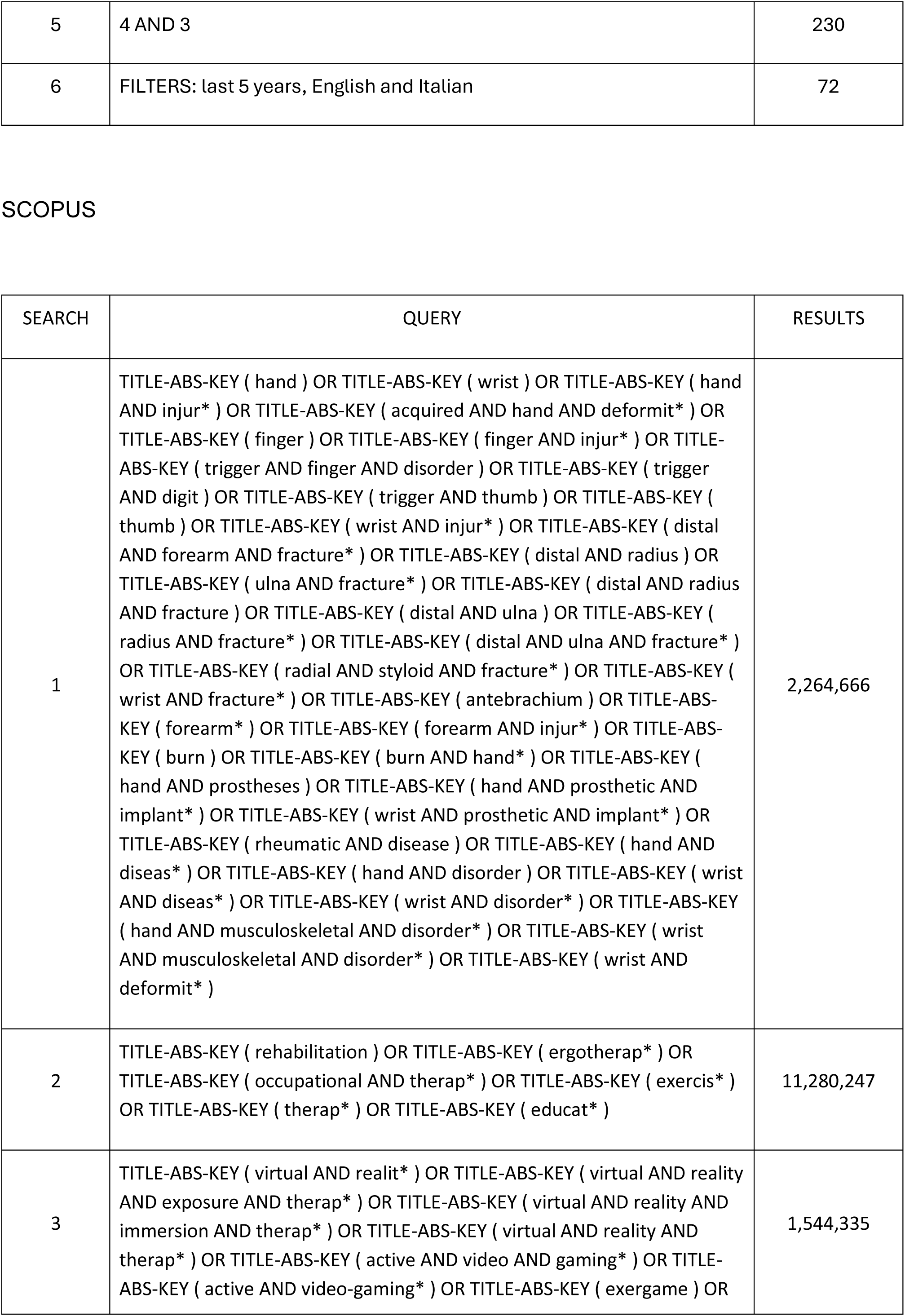
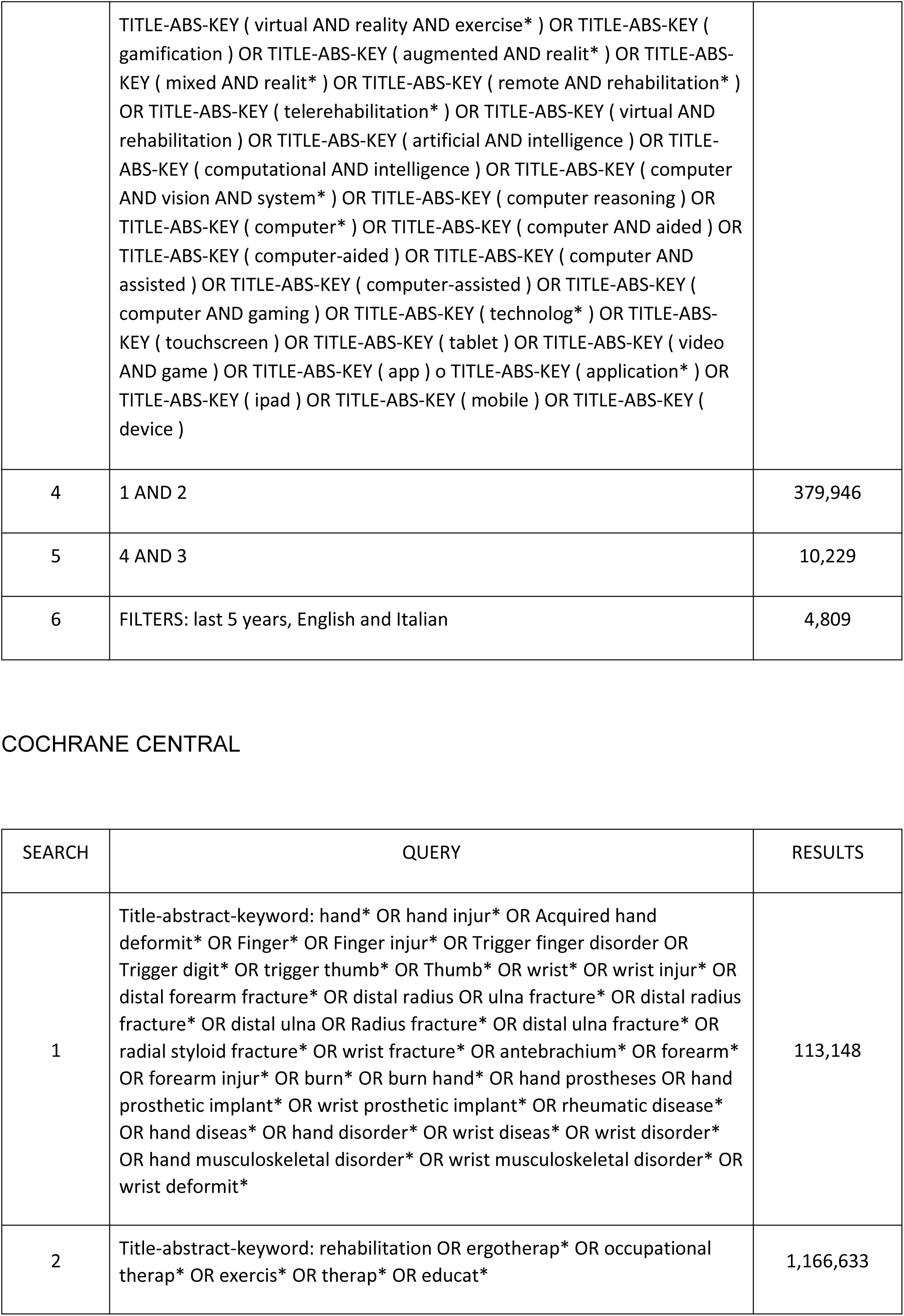
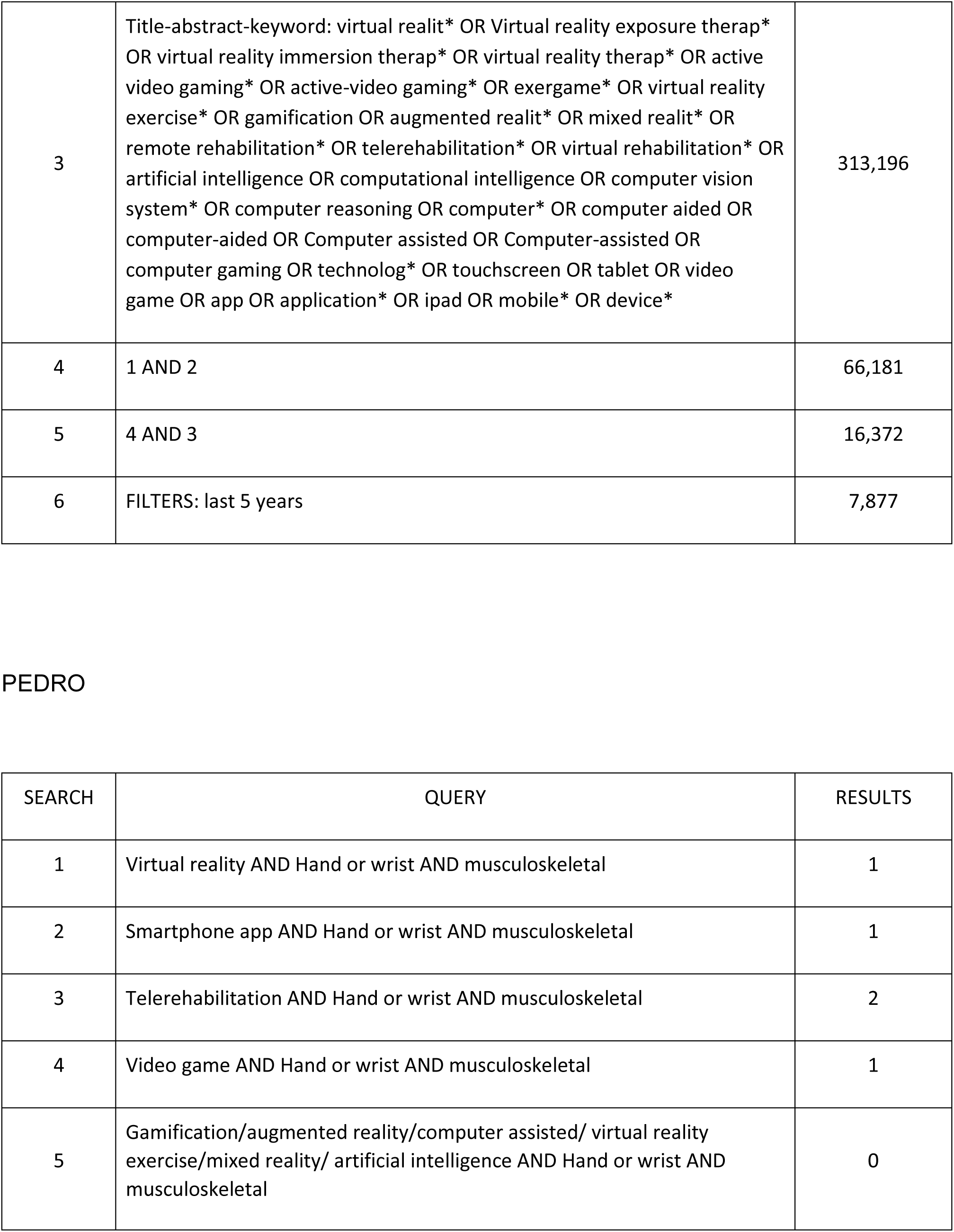
research strategies for each database.

All articles written since 2019 will be included in this scoping review. Articles written in languages other than English or Italian will not be included. Zotero software (Roy Rosenzweig Center for History and New Media Elsevier Incorporation, Fairfax, Virginia, USA) will be used for citation management and PRISMA-S for reporting the results of the study. The search started on 8th December 2024.

### Study selection

After applying the inclusion criteria and once the bibliographic search is complete, all identified results will be collected and entered into the *Rayyan.ai* Software and duplicates will be removed. Subsequently, the abstracts of the emerged articles will be examined and, if this part is missing, the entire paper will be read directly. If the entire text of the selected article is not present, the authors will be contacted for no more than three attempts. Two reviewers will carry out the article selection process. A third reviewer will resolve any discussions. During this phase, all relevant information will be entered into *Microsoft Excel* (Microsoft Corporation, Redmond, WA, USA) and presented in Preferred Reporting Items for Systematic Reviews and Meta-analyses flow diagram.

Any exclusion of full-text articles will be reported in this Scoping Review with relative reasons.

## Data Extraction

The data extraction sheet will be designed to systematically collect key information from each included article. The following data points will be extracted:

- Basic Information: authors, year of publication, country of study.
- Study Design: Type of study (e.g., randomized controlled trial, observational study), sample size.
- Participant Characteristics: Age, diagnosis.
- Intervention Details: Type of VR or technologic device used, mode of application, and duration of the treatment.
- Outcomes: Primary and secondary outcomes, including both quantitative and qualitative data.
- Limitations: Declared or potential biases and other limitations identified in the study.

Data collection will be performed using Microsoft Excel or equivalent software. To ensure accuracy, two reviewers will independently extract the data. Any discrepancies will be resolved through discussion or by consulting a third reviewer. The extracted data will form the basis for addressing the review questions and identifying gaps in the literature.

The information gained will be included and discussed in this Scoping Review. A background analysis will be proposed in this study regarding details of participants, mode of administration, clinical repeatability, links between articles and other information useful in resolving the Review Questions.

If any information relevant to this study is unclear or missing, the authors of these articles will be contacted three times. If this information is not provided, the missing information will be stated as ‘*Not available’* and the doubtful information as *‘Unclear’*.

## Synthesis and Presentation of Results

The information gained from the articles will be presented in a Prisma Flow Diagram to describe the article selection process. A PRISMA-compliant flowchart will be created to illustrate the process of study selection, from initial search to final inclusion. The flowchart will include the following steps:

Identification:

- Number of records identified through database searches (e.g., PubMed, Cochrane Central, Scopus, Pedro, CINAHL Complete)
- Number of records identified from grey literature sources (e.g., Google Scholar).

Screening:

- Number of records after removing duplicates.
- Number of records excluded based on title and abstract screening.

Eligibility:

- Number of full-text articles assessed for eligibility.
- Number of full-text articles excluded, with reasons (e.g., wrong population, intervention, or outcomes).

Inclusion:

- Number of studies included in the final scoping review.

The flowchart will visually represent these steps, ensuring transparency in the decision-making process for article inclusion and exclusion.

Subsequently, the data from each study will be presented in tables and described in a discursive manner to answer the questions and subject matter of this Review. The framing of the themes will be used to produce a thematic synthesis. This will be related to the key concepts and themes relevant to the research questions and according to the subgroups (e.g. different type of virtual reality or technological devices, or different diagnosis) and others that may emerge. If necessary, the approach used to summarise and present the results may be modified during the review process and based on the selected articles.

## Ethics and Dissemination

No Ethical approvation will be necessary. The results will be disseminated in peer-review journals also with the aim of providing recommendations for possible future research.

## Data Availability

all data produced in the present work are coneined in the manuscript

## Acknowledgements

None

